# Increasing the resolution of malaria early warning systems for use by local health actors

**DOI:** 10.1101/2024.04.17.24305948

**Authors:** Michelle V Evans, Felana A Ihantamalala, Mauricianot Randriamihaja, Vincent Herbreteau, Christophe Révillion, Thibault Catry, Eric Delaitre, Matthew H Bonds, Benjamin Roche, Ezra Mitsinjoniala, Fiainamirindra A Ralaivavikoa, Bénédicte Razafinjato, Oméga Raobela, Andres Garchitorena

## Abstract

**Background:** The increasing availability of electronic health system data and remotely-sensed environmental variables has led to the emergence of statistical models capable of producing malaria forecasts. Many of these models have been operationalized into malaria early warning systems (MEWSs), which provide predictions of malaria dynamics several months in advance at national and regional levels. However, MEWSs do not generally produce predictions at the village-level, the operational scale of community health systems and the first point of contact for the majority of rural populations in malaria-endemic countries.

**Methods:** We developed a hyper-local MEWS for use within a health-system strengthening intervention in rural Madagascar. It combined bias-corrected, village-level case notification data with remotely sensed environmental variables at spatial scales as fine as a 10m resolution. A spatio-temporal hierarchical generalized linear regression model was trained on monthly malaria case data from 195 communities from 2017-2020 and evaluated via cross-validation. The model was then integrated into an automated workflow with environmental data updated monthly to create a continuously updating MEWS capable of predicting malaria cases up to three months in advance at the village-level. Predictions were transformed into indicators relevant to health system actors by estimating the quantities of medical supplies required at each health clinic and the number of cases remaining untreated at the community level.

**Results:** The statistical model was able to accurately reproduce village-level case data, performing nearly five times as well as a null model during cross-validation. The dynamic environmental variables, particularly those associated with standing water and rice field dynamics, were strongly associated with malaria incidence, allowing the model to accurately predict future incidence rates. When compared to historical stock data, the MEWS predicted stock requirements within 50 units of reported stock requirements 68% of the time.

**Conclusion:** We demonstrate the feasibility of developing an automatic, hyper-local MEWS leveraging remotely-sensed environmental data at fine spatial scales. As health system data become increasingly digitized, this method can be easily applied to other regions and be updated with near real-time health data to further increase performance.

## BACKGROUND

Data systems play a key role in malaria control initiatives. Indeed, malaria surveillance is one of three pillars of the World Health Organization’s (WHO) Global Technical Strategy for Malaria 2016-2030 (World Health Organization 2021). The strategy stresses the need to strengthen local health management information systems (HMISs) to better track progress towards elimination and heterogeneity within a country. In addition to surveillance, malaria early warning systems (MEWSs), which use statistical and mathematical models to forecast malaria dynamics up to several months in advance as a function of environmental variables, can aid health systems in preventing malaria outbreaks and improving system reactivity (Zinszer et al. 2012, Hemingway et al. 2016). Relevant environmental variables, such as temperature, precipitation, and vegetation indices, can be derived from satellite imagery, whose resolution, frequency, and accessibility are continuously improving (Wimberly et al. 2021). With the evolving capabilities of health and environmental data systems, it is increasingly feasible to link these two data systems and create disease forecasts. However, while several MEWSs have been developed at global, regional, and national scales, the routine integration of environmental data in MEWSs remains rare (Hussain-Alkhateeb et al. 2021).

The spatial scale at which MEWSs are developed determines the potential users of the tool for decision-making. While current MEWSs can effectively inform international organizations and national program managers, few MEWSs have been developed at local scales (< 1km^2^) relevant for operational use by health actors implementing malaria control activities within a health district. Increasing the resolution of MEWSs could allow district managers and medical teams to adapt interventions to the village or local level, such as hotspot targeting, last mile delivery, and community health worker (CHW) programs. Malaria hotspots (zones with consistently high incidence rates) have been proposed as an appropriate unit to target with vector control or human health interventions such as mass drug administration (Bousema et al. 2012), although the ability of these interventions to impact regions outside of hotspots is limited (Bousema et al. 2016, Stresman et al. 2019).

Last mile delivery interventions support the management and distribution of medical stocks such as antimalarials and bed nets at the lowest scale of the health system, often the CHW or even households delivered door-to-door (USAID 2023). By delivering medical products and services to remote populations, these programs aim to remove geographic barriers to prevention and treatment of malaria within a medically-relevant timeframe. CHW programs provide basic maternal and child care within local communities of several hundred to several thousand people (Perry 2020). CHWs traditionally diagnose and treat malaria in children under 5 years of age, and recent pilot programs have demonstrated the success of expanding responsibilities to include novel malaria interventions, such as the provision of intermittent preventive treatment to pregnant women (Pons-Duran et al. 2021) or proactive screening and treatment (Ratovoson et al. 2022). These successes of community-targeted programs have prompted calls for the increased development of digital health tools for programs implemented at local scales, especially in sub-Saharan Africa (Holst et al. 2020, Owoyemi et al. 2022).

In order for a MEWS to be usable by programs at the local-scale, it must not only provide predictions at a relevant spatial scale, but it should also demonstrate low latency and contextual relevance. This presents challenges as HMIS data are rarely reported at the scale of individual villages or communities, and when they are, tend to suffer from substantial data quality issues and biases (Noor et al. 2003, Ohrt et al. 2015, Garchitorena et al. 2021). In addition, the predictive variables used in the MEWS statistical model must also be at finer spatial scales than typically available, necessitating the pre- and post-processing of satellite imagery in continuous time (Wimberly et al. 2022). Finally, outputs of a MEWS should be made contextually relevant by integrating predictions with existing HMIS data, such as historical case burdens, information on diagnostic and treatment supplies, and ongoing programs. In consideration of these challenges, we developed SMALLER (‘Surveillance and control of Malaria At the Local Level using E-health platfoRms’), a hyper-local MEWS specifically for use in the context of a health-system strengthening intervention in a rural district of Madagascar.

## METHODS

We collected HMIS data and socio-environmental variables at the spatial scale of community health catchments, ensuring data quality by treating HMIS data with a zero-adjusted, gravity model estimator (Evans et al. 2023) and correcting satellite imagery with multiple treatment and gap filling processes, allowing us to retain the original scales of the data. We paired these data sources with a geographic information system of all the residential areas and rice fields in the district collected via a participatory mapping project (Ihantamalala et al. 2020), allowing us to extract data in zones where malaria transmission is likely occurring.

Geostatistical models were trained using a Bayesian framework that allowed for spatio- temporal random effect structures to leverage information across community health catchments. Finally, we integrated the full workflow into a web application architecture that updates predictions continuously and provides information to local health actors in formats directly applicable to decision-making processes within the district (Fig. 1).

**Figure 1.**
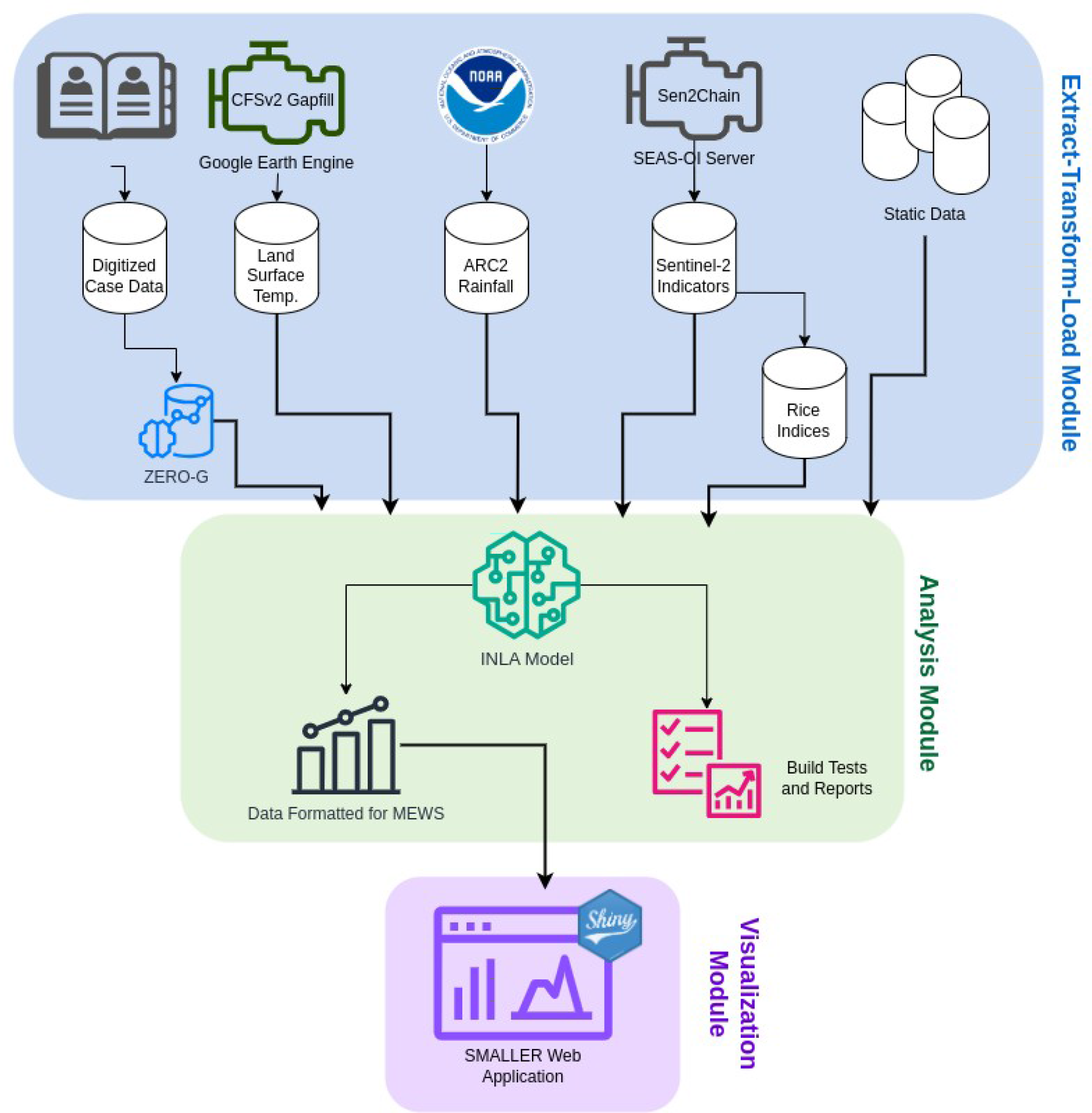
Schema of SMALLER MEWS web application architecture. The workflow is divided into three modules: Extract-Transform-Load, Analysis, and Visualization. The full workflow is updated on a monthly basis via a semi-automated process using the `targets` package.

### Study Area

Madagascar is one of the few countries that has seen an increase in malaria burdens since the beginning of global elimination efforts in the early 2000s (Howes et al. 2016) The southeastern part of Madagascar experiences unimodal seasonal cycles of malaria (Nguyen et al. 2020), with overall higher prevalence rates than the central plateau regions (Ihantamalala et al. 2018, Arambepola et al. 2020, Rice et al. 2021). Ifanadiana is a rural health district located in the Vatovavy region of southeastern Madagascar. The district’s population is approximately 200,000 people, the majority of whom live in rural, isolated villages over 1 hour from a health center (Ihantamalala et al. 2020). The district is divided into 15 communes and 195 fokontany (the smallest administrative unit comprising one or several villages amounting to about 1000 individuals, which represents the community health catchment). Each commune contains at least one major primary health center (PHC), and six of the larger communes contain a second, basic primary health center that do not have medical doctors and provide more limited services (Fig. S4.1). Beginning in 2014, Ifanadiana has benefitted from a health system strengthening (HSS) intervention at all levels of the health system, from community health to the regional hospital, via a partnership between the Madagascar Ministry of Public Health (MMoPH) and the non-governmental organization Pivot (Cordier et al. 2020).

Ifanadiana contains a variety of ecoregions, including a protected tropical rainforest in the west and warmer, humid zones near the eastern coast. There is an east-west elevational gradient from an altitude of 1400m in the west to 100m in the east. The dominant land covers are savanna and agricultural land for rice production. This diversity of ecoregions and climates translates into spatio-temporal heterogeneities in malaria burden in the district (Hyde et al. 2021, Pourtois et al. 2023).

### Data Collection

#### Health Data

The monthly number of malaria cases per community were collected from consultation registries at all primary health centers (PHCs) across the district, from January 2017- December 2020. Handwritten registries from each PHC were digitized, with each de-identified patient geolocated to the precision of a fokontany. The number of malaria cases, as confirmed by rapid diagnostic test (RDT), were aggregated by month for children under 5 years old, children aged 5 - 14, and adults aged 15 years and above. Because these data are collected at PHCs, they are passive surveillance data and contain primarily symptomatic cases. These raw data were adjusted for underascertainment due to spatial bias in healthcare access using the ZERO-G method (Evans et al. 2023), which accounts for whether the PHC fell within the initial Pivot service catchment, whether point-of-care user fees had been removed for the PHC, the number of staff at the PHC, the level of the services, and the distance from the PHC to the district office. We also included additional identifiers of false-zeros based on the geographic coverage of the HSS intervention (see Supplemental Materials for more details). The ZERO-G method was applied seperately to each age class. The final data used in the model were the ZERO-G adjusted monthly case counts, aggregated across all ages.

Information on historical quantities of malaria diagnostics and treatment were used to validate model predictions and provide additional context for decision makers in the web application. These data were provided by the MMoPH at the monthly level for all major PHC in the district beginning in 2017 and are updated continuously. This includes the number of febrile patients seen at the facility, the number of febrile patients tested via RDT, the number of malaria positive RDTs, and the number of RDT-positive patients treated with Artemisinin-based combination treatments (ACTs).

#### Environmental Indicators

The surrounding environment can greatly influence the ecology of *Anopheles* mosquitoes, and therefore malaria dynamics themselves. We therefore collected data that described environmental dynamics, such as landcover, hydrology, and vegetation from the zones surrounding residential areas. We located major residential zones using a comprehensive dataset of the district of Ifanadiana collected via participatory mapping and available on OpenStreetMap (Ihantamalala et al. 2020). This dataset includes over 25,000 km of roads, tracks, and footpaths, 20,000 rice fields, 5000 residential areas, and 100,000 buildings. The dataset used for this study was accessed on Nov 30 2021. From this dataset, we identified the four primary villages of each fokontany, determined by the number of buildings and field-based knowledge of communities. These zones contained on average 40% of buildings in a fokontany and 60% of all buildings within residential zones, with the remaining buildings usually standalone structures such as small houses or shelters used during the agriculture season, rather than residences. We focused on the environment within 1 km of these residential zones, assuming that mosquito’s flight distances were limited, following previous work in this region (Arisco et al. 2020). In addition, we identified the four largest rice fields in close proximity to these villages, creating a dataset of potential larval habitat closest to human settlements. This resulted in a total of 775 primary villages with a 1 km buffer and 769 major rice fields that were used to extract environmental variables (Fig. 2).

**Figure 2.**
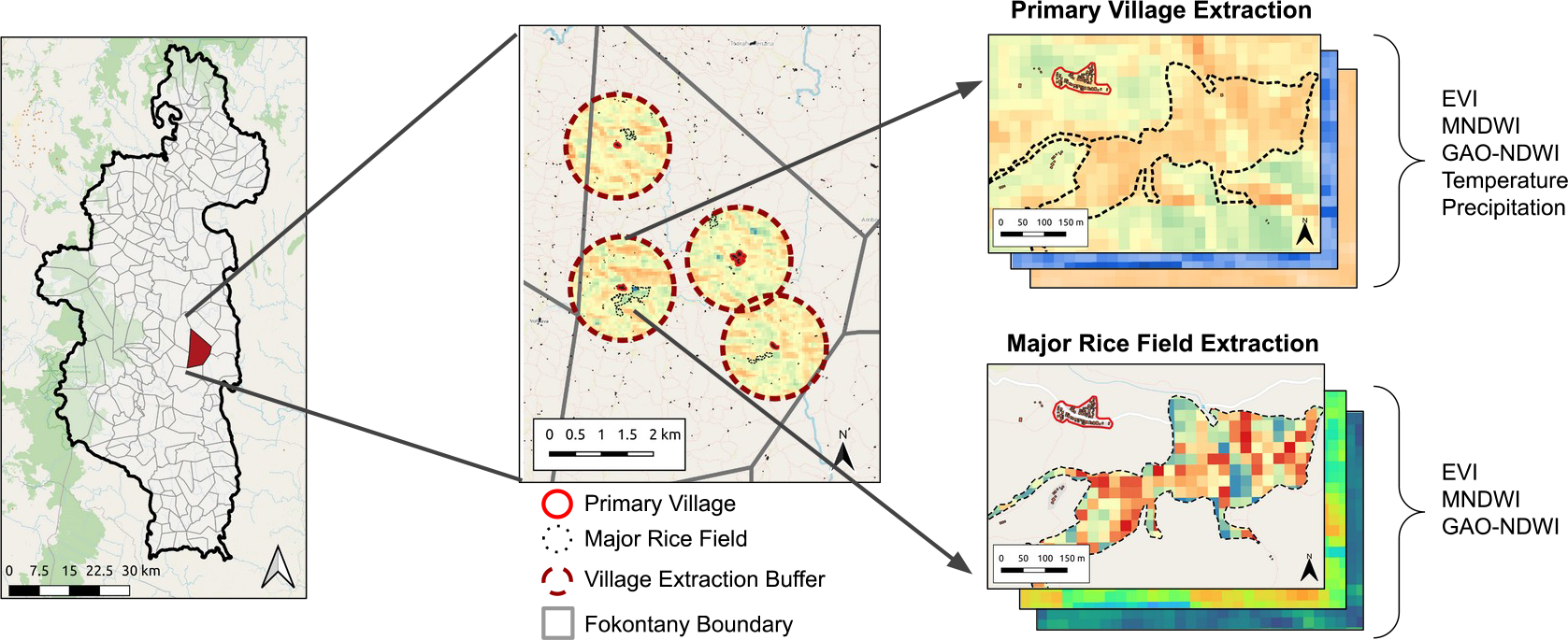
Process of extracting environmental indicators at a fine-scale from satellite imagery for major village and rice field extraction zones. Indicators at the primary village level are extracted using a 1km buffer surrounding the village (red-dashed line), while indicators at the rice field level are extracted within the boundaries of the major rice fields (black dashed line).

Landcover variables were calculated from a dataset derived from OpenStreetMap and Sentinel-2 satellite imagery, as described in Evans et al. (2021). This dataset classified landcover into residential, rice fied, savanna, forest, and open water at a 10m resolution. We estimated the proportion of landcover that was a rice field within the 1 km buffer of the primary villages. We aggregated this value to the level of the fokontany by calculating a mean weighted by the population size of each primary village, multiplying the values by the proportion of the buildings in the residential area out of all buildings in all four residential areas (i.e. a building-weighted mean).

We derived two metrics representing spatial patterns in hydrology and standing water from the SRTM 30-m digital elevation model (DEM). Topographic wetness indices (TWIs) estimate predicted water accumulation as a function of an area’s upstream catchment area and slope, and have been shown to predict malaria risk in regions with highly varied topologies (Cohen et al. 2010, Platt et al. 2018). We estimated the TWI as 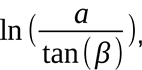, where *a* is the surface catchment area of that pixel and *β* is the local surface topographic slope. We calculated the spatial mean of the TWI at each primary village and major rice field within a fokontany and calculated the mean TWI at the village and rice field levels within a fokontany by taking a building-weighted and standard mean, respectively. From the same DEM, we identified cells that served as sinks, or pixels with no outflow where water is likely to accumulate. We extracted the proportion of pixels that were sinks within a 1 km buffer of each primary village and used a building-weighted mean to calculate a fokontany-level value. All topographic analyses were done using GRASS GIS (GRASS Development Team 2019) via the `rgrass` package in R (Bivand 2023).

Vegetation indices were derived from Sentinel-2 satellite imagery. These optical satellites have a revisit time of 5 days and a 10m spatial resolution. We used the Sen2Chain Python tool (Mouquet et al. 2020) to download and process the Sentinel-2 imagery to level L2A before calculating three radiometric indices: the Enhanced Vegetation Index (EVI) (Huete et al. 1997), the Modified Normalized Difference Water Index (MNDWI) (Xu 2006), and Gao’s Normalized Difference Water Index (GAO-NDWI) (Gao 1996). Each of these indices represents different aspects of rice field land cover relevant for *Anopheles* habitat.

EVI represents vegetation health and vigor, MNDWI identifies areas of open water, and NDWI-GAO estimates moisture in the vegetation. Images with greater than 25% cloud cover were removed and indices were extracted at the primary village 1km buffer and major rice fields as described above (Fig. 2). Each extraction zone (e.g. primary village or major rice field) was less than 3 km^2^ and therefore very susceptible to measurement error and random fluctuations. To reduce this stochastic noise and interpolate measurements for dates with high cloud cover, we smoothed the time series for each zone by applying a cubic-spline to the series, using leave-one-out cross-validation to select the optimal number of degrees of freedom. This cubic-spline was then used to predict values on a weekly frequency for each index. Monthly means were calculated from these weekly values for each fokontany as follows: village-level indicators were aggregated by the building-weighted mean described above, and rice field indicators were estimated as the mean value of all major rice fields in the fokontany. The final result was a monthly time series at the fokontany level for the three environmental indices at both the village and rice field zones.

We used principal component analysis to create indices of rice field dynamics from EVI, MNDWI, and GAO-NDWI extracted from rice fields zones. First, we estimated the difference in each index at the rice field level from the index extracted at a buffer of 1 km surrounding village zones (Δvill-rice) to categorize dynamics specific to rice field environments. Second, we transformed the indice extracted at rice fields (EVI, MNDWI, and GAO-NDWI) into seasonal anomalies by standardizing each index within each calendar month (e.g. January, February, etc.) following Kaul et al. (2018). This seasonal anomaly represents how conditions differed in that year compared to the same calendar month over the long-term dataset. Third, we included the original three environmental indices extracted at the major rice field zones. We used all three forms of the three indicators (Δvill-rice, seasonal anomalies, and original mean; 9 variables total) in the PCA, after having centered and scaled each one. We then selected the first three components, which contained over 74% of the overall variance, to represent three indices of rice field dynamics (Table S1.1, Table S1.2, Table S1.3). Rice Index 1 was strongly influenced by all three forms of the MNDWI indicator, and was positively associated with the amount of standing water in rice fields, representing wetter periods. Rice Index 2 was more strongly associated with EVI and NDWI-GAO and represented the vegetation dynamics of the rice fields, with higher values associated with greener vegetation in rice fields compared to the surrounding village. Rice Index 3 was most strongly associated with anomalies in the vegetation indices and represented anomalies in vegetation phenology (specifically increased vegetation) and the timing of the agricultural season.

#### Climate Data

Meteorological stations are rare in Madagascar. Satellite-derived climate data at resolutions less than 5km are equally limited and, when present, suffer from cloud obstruction, particularly in the humid southeastern region. One solution is to aggregate data to a coarser resolution or over multiple time periods, but this results in spatial scales too coarse to accurately represent hyper-local conditions. We considered this lack of fine-scale, accurate data in our choice of data sources and processing of both temperature and precipitation data. We estimated land surface temperature (LST) via a gap-filling algorithm that combines climatology from fine-scale MODIS imagery with modeled surface temperature from the Climate Forecast System Version 2 (CFSv2) to create daily estimates of LST at a 1 km resolution (Shiff et al. 2021). This method has been validated globally and performs well in this region of Madagascar, with a RMSE less than 2°C. We specifically used the MODIS Aqua Daily Land Surface Temperature, which was averaged to a monthly value after gap- filling. This represents an improvement in resolution over the 8km resolution corrected data available directly via MODIS at the monthly level. We obtained precipitation data from the NOAA Africa Rainfall Climatology v2 (ARC2) daily precipitation dataset via the `rnoaa` package in R (Chamberlain and Hocking 2023). We selected this dataset due to its high reported accuracy for this region of Madagascar (Ollivier et al. 2023). We summed the daily precipitation by month to obtain the total monthly precipitation for each 0.1 x 0.1 degree (∼ 10 km) pixel. Both climate variables were extracted using a primary village 1km buffer and aggregated to the fokontany-level via a building-weighted mean, as described above.

#### Socio-demographic data

Socio-demographic data were collected from 2014-2021 via the IHOPE cohort, a longitudinal survey based on the Demographic and Health Surveys, conducted in about 1600 households of Ifanadiana district distributed across 80 spatial clusters (Miller et al. 2018).

Briefly, a two-stage sampling design was used to sample 40 clusters at random with each of two strata, the initial HSS intervention catchment and the rest of the district. Twenty households per cluster were then randomly selected to be surveyed. Further details on the IHOPE longitudinal survey can be found in Miller et al. (2017, 2018). The IHOPE cohort collects information on household-level socio-demographic, health, and socio-economic indicators. We included data on household wealth scores in this study, extracted to the fokontany level following Evans et al. (2022).

In addition to household-level demographic data, we included variables representing the geography of each fokontany, specifically the distribution of houses and the distance to primary health centers. Residential areas are clustered within Ifanadiana district and a fokontany-wide estimate of building density does not accurately represent the population density experienced by an individual. We therefore calculated a relative building density by calculating the density of buildings within 100m of a building for all buildings within each fokontany. We calculated the median of this value to obtain fokontany-level estimates. The distance to a primary health center was estimated for each building over the full transport network using the Open Source Routing Machine (OSRM) routing algorithm via the `osrm` package in R (Giraud 2022), and the average distance was calculated for each fokontany.

#### Health Intervention Data

The study period intersected with a time of on-going health system interventions known to impact malaria dynamics. Over the past decade, Madagascar has conducted a mass long-lasting insecticide-treated bed net (LLIN) distribution every three years beginning in October 2015. The effect of bed nets decreases over time due to waning bioefficacy and functional integrity (Ngonghala et al. 2014, Randriamaherijaona et al. 2017). We therefore included a variable in the model to represent this waning over time via the number of months since the most recent bed net distribution. From October 2019 thru December 2021, a pilot proactive community care intervention was implemented in one commune in the district, increasing treatment rates of malaria by nearly 40% (Razafinjato et al. 2024). We accounted for this heterogeneity in treatment rates caused by interventions by including a binary variable for months and fokontany when the proactive care intervention was in place. Finally, although the data were already adjusted for geographic bias in health-seeking behaviors via ZERO-G, an artifact of this bias remained. We therefore included the distance to the nearest PHC in the model, allowing for a non-linear relationship via a penalized smoothing spline.

### Statistical Model

We implemented a Bayesian spatio-temporal model via an Integrated Nested Laplace Approximation (INLA) model that included hierarchical random effects (Rue et al. 2009). Bayesian hierarchical models are particularly useful for analyzing spatio-temporal data because of their ability to leverage random effects across space and time to account for spatio-temporal correlation and more accurately estimate random effect coefficients when data are of low quality or missing. We accounted for the spatio-temporal covariance in our data in two ways. First, we included a cyclical temporal term by month of the year via a first order random walk, estimated for each commune. Second, we included spatial covariance by fokontany via the Besag, York, and Mollie spatial model (Besag et al. 1991, Morris et al. 2019), which includes an unstructured random effect for each fokontany in addition to a Besag model for the spatial structure. Predictor variables were inspected for normality and log-transformed when necessary, then scaled and centered to aid with model convergence. Dynamic variables were lagged by 3 months to account for delays in the effect of environmental and climatic variables on malaria transmission and to allow for predicting malaria trends into the future. We conducted a supplementary analysis to explore including non-linear effects of predictor variables via penalized splines, but found that it performed similarly to a model including only linear effects while requiring a significantly longer computation time (details reported in the Supplementary Materials). We therefore used the more parsimonious linear model. This resulted in a total of fourteen covariates in the model, six of which are lagged, dynamic variables updated monthly (Fig. S4.2, Table S5.1). The INLA model was fit to monthly case data at the fokontany level via a zero-inflated negative binomial distribution using a log-link, with population as an offset. The model was trained on the number of malaria cases per fokontany from January 2017 thru December 2020 and used to predict future disease incidence for the MEWS.

We performed out-of-sample prediction tests across both space and time to assess our model’s predictive capabilities via leave-one-out cross-validation. Spatial out-of-sample assessment was done by commune, leaving one commune out of model training and predicting incidence in that commune, resulting in 15 separate tests. Temporal out-of-sample assessment was done by year, where each sample omitted from model training corresponded to a year of data from 2017-2020. We assessed the model’s predictive ability via the IQR of the absolute error, which is more robust to outliers than the RMSE, and the Spearman’s correlation coefficient (ρ) between the predicted and observed incidence rates.

A motivation for the creation of this dashboard was the high frequency of disruptions to diagnostic and medical stocks (“stock-outs”) observed in malaria-endemic regions of Madagascar, and the need for better guidance to plan future stock use. We therefore further validated the model retrospectively by comparing predicted ACT needs (i.e. predicted number of malaria cases expected to seek care) to reported ACT needs at each PHC (i.e. number of positive malaria cases requiring treatment). To do this, we first back-calculated the number of cases per fokontany expected to seek care at a PHC by rescaling the total number of cases per fokontany. We assumed a binomial probability of observation of each case at a PHC equal to the sampling intensity estimated via the ZERO-G method. The cases that did not seek care were assumed to remain at the community health catchment. We then predicted the proportion of cases attending each PHC from each fokontany based on the historical distribution of consultations from each fokontany to each PHC from 2018-2019. We chose this subset of the data in order to use the most recent and most complete consultation data that did not suffer from bias due to the COVID-19 pandemic which began in 2020.

These numbers were then aggregated to the level of the PHC, providing the number of malaria cases predicted to seek care at each PHC, or the predicted ACT requirement. We validated this method of back-calculation by comparing predicted ACT requirements from the SMALLER MEWS to reported ACT requirements for 14 major PHCs from January 2017 - December 2020 by estimating the absolute error and Spearman’s correlation coefficient between the two datasets.

### Automating the Workflow for a MEWS Web Application

We automated the remote sensing and statistical model workflow for a MEWS web application via the `targets` package (Landau 2021), which creates a Make-like pipeline for R scripts. The use of a Make-like workflow is especially beneficial for deploying a MEWS in a resource-limited setting because only those data sources and tasks that are not up to date are rerun in the monthly update, conserving computational and network resources. The `targets` pipeline “backend” is linked to an R shiny “frontend”, which contains the web application user interface (Fig. 1). The workflow updates monthly, collecting new environmental variables and creating updated forecasts that are then available online for use by local health actors.

The `targets` pipeline contains the Extract-Transform-Load and Analysis modules of the application (Fig. 1). Static data are loaded into the project one time, pre-processed and formatted for the model. Dynamic data are then updated monthly on the tenth day of the month, to allow for latency in data collection, before being combined with the static data to use in prediction. We use a combination of tools to collect this data semi-automatically.

Google Earth Engine scripts, which process temperature data, are manually run via the Online Code Platform, and processed data are added to the project, where they are tracked via the `targets` workflow. Sentinel-2 indices are updated monthly via the Sen2Chain tool implemented on servers hosted at SEAS-OI Station at Université de la Réunion, where the data are then provided through HTTP. The data source url is added to the `targets` workflow, which then observes the HTTP file for changes. If a change is made to the file, the new data are automatically included in the next update. Because the `targets` workflow tracks the downstream flow of data, when the raw index data are updated, all downstream features derived from this, such as the rice indices, are also updated. The workflow is semi- manual, in that the automatic pipeline is run under supervision, so that the input data and resulting predictions can be validated by a subject expert before being made publicly available. This is done via a `quarto` document that automatically renders after the workflow is completed, providing information on the environmental input data and output predictions.

## RESULTS

### Malaria Case Data

In total, there were 107,739 reported malaria cases in Ifanadiana district from January 2017 - December 2020. Missingness of data (*i.e.*, registers not available) was 2.21%, or 207 of 9360 total month by fokontany samples. After applying the ZERO-G method, which adjusts for underascertainment due to spatial bias in healthcare access, the estimated total number of symptomatic malaria cases was 377,211, with a median annual incidence per fokontany of 357 cases per 1000 individuals (95% CI : 2.81 - 1699). The highest incidence was observed between the months of November - April, ranging from 37.2 – 77.1 cases per 1000 individuals per month. The mean annual incidence was 1626 cases per 1000 in the 25% of fokontany experiencing the highest incidence rates and 280 cases per 1000 in the 25% of fokontany experiencing the lowest incidence rates.

### Model Performance and Results

The INLA model was able to accurately reproduce community-level malaria case data. When applied to the full dataset, it achieved a median absolute error of 16.99 cases per 1000 individuals per month (IQR = 7.30 – 38.93), equivalent to 43% of the mean monthly adjusted incidence rate. The predicted and oberved incidence rates were significantly positively correlated (Spearman’s p = 0.647, p-value <0.0001). Similarly, the model performed well when assessed via cross-validation across space and time (Table 1). Predictive ability was similar across communes, with the exception of one commune in the south of the district (Fig. S4.3), where accuracy was much lower than in the other 14 communes. The model performed well at predicting incidence in out-of-sample years. In particular, the full model performed better on out-of-sample data in 2020 than on in-sample data, evidence of the models’ ability to forecast forward in time (Fig. S4.4).

**Table 1.** Performance metrics of the predictive model when assessed via leave-one-out cross-validation across time and space. The null model corresponds to a model that only estimates a district level mean without any covariates. Values are the mean performance metrics with the range of metrics in parentheses. Median absolute error (AE) represents the median absolute difference between model predictions and the true values, with lower values representing better model performance. Spearman’s ρ represents the correlation between the model predictions and the true values, with values closer to one representing better model performance.

|  | Temporal cross-validation<br>(by year) |  | Spatial cross-validation<br>(by commune) |  |
| --- | --- | --- | --- | --- |
|  | Null Model | Full Model | Null Model | Full Model |
| In-Sample<br>Median AE | 36.23 (35.64 - 36.5) | 15.93 (15.46 - 16.69) | 36.24 (35.22 - 37.82) | 15.93 (15.35 - 17.03) |
| Out-of-Sample<br>Median AE | 36.26 (35.6 - 38.13) | 15.91 (14.03 - 17.25) | 40.39 (22.75 - 67.25) | 18.55 (9.53 - 48.72) |
| In-Sample<br>Spearman's $\rho$ | 0.10 (0.09 - 0.12) | 0.66 (0.63 - 0.69) | 0.10 (0.07 - 0.13) | 0.66 (0.65 - 0.68) |
| Out-of-Sample<br>Spearman's $\rho$ | 0.10 (0.05 - 0.13) | 0.66 (0.54 - 0.74) | 0.05 (-0.12 - 0.27) | 0.67 (0.41 - 0.84) |

SMALLER MEWS predicted stock requirements by combining predicted malaria incidence rates with estimated health-care seeking behaviors, which were compared with reported stock requirements. Reporting of historical stock requirements was incomplete, with 12 of 14 PHCs missing the quantity of ACTs required (e.g. the number of positive RDTs) during at least one month. However, only 3 PHCs were missing more than 5 months of data over the 48 month period. The reported ACT requirement in the district ranged from 15,624 to 38,154 annually, with needs increasing steadily over time from 2017 - 2020 (Fig. 3). In contrast, the predicted ACT requirement ranged from 20,736 to 28,399 per year over the same time period. The median absolute error between predicted and reported ACT requirements was 20 units (IQR = 9 - 70), equaling an error of approximately 20% of the mean number of monthly cases received at a PHC. However, the dataset of reported ACT requirements was strongly influenced by outliers. When assessed via a Spearman’s correlation test, which is more robust to outliers, the predicted ACT requirements correlated strongly with the reported ACT requirements (Spearman’s p = 0.849, p-value<0.001). The SMALLER MEWS estimated the quantity of ACT required within 50 units of the reported requirement for 68% of the month and PHC samples. It did not show a strong bias for under- or over-estimation, overestimating the requirement for 44.2% of the samples and underestimating the requirement for 55.8% of the samples. Performance was dependent on the PHC, with the average annual difference in predicted vs. reported stock needs ranging from -73.36% to +15.25 % of the actual need across all 14 PHCs (Fig. 3). For all PHCs, there were months with very high reported ACT requirements that were underestimated by the SMALLER MEWS, particularly in 2020 (Fig. 3).

**Figure 3.**
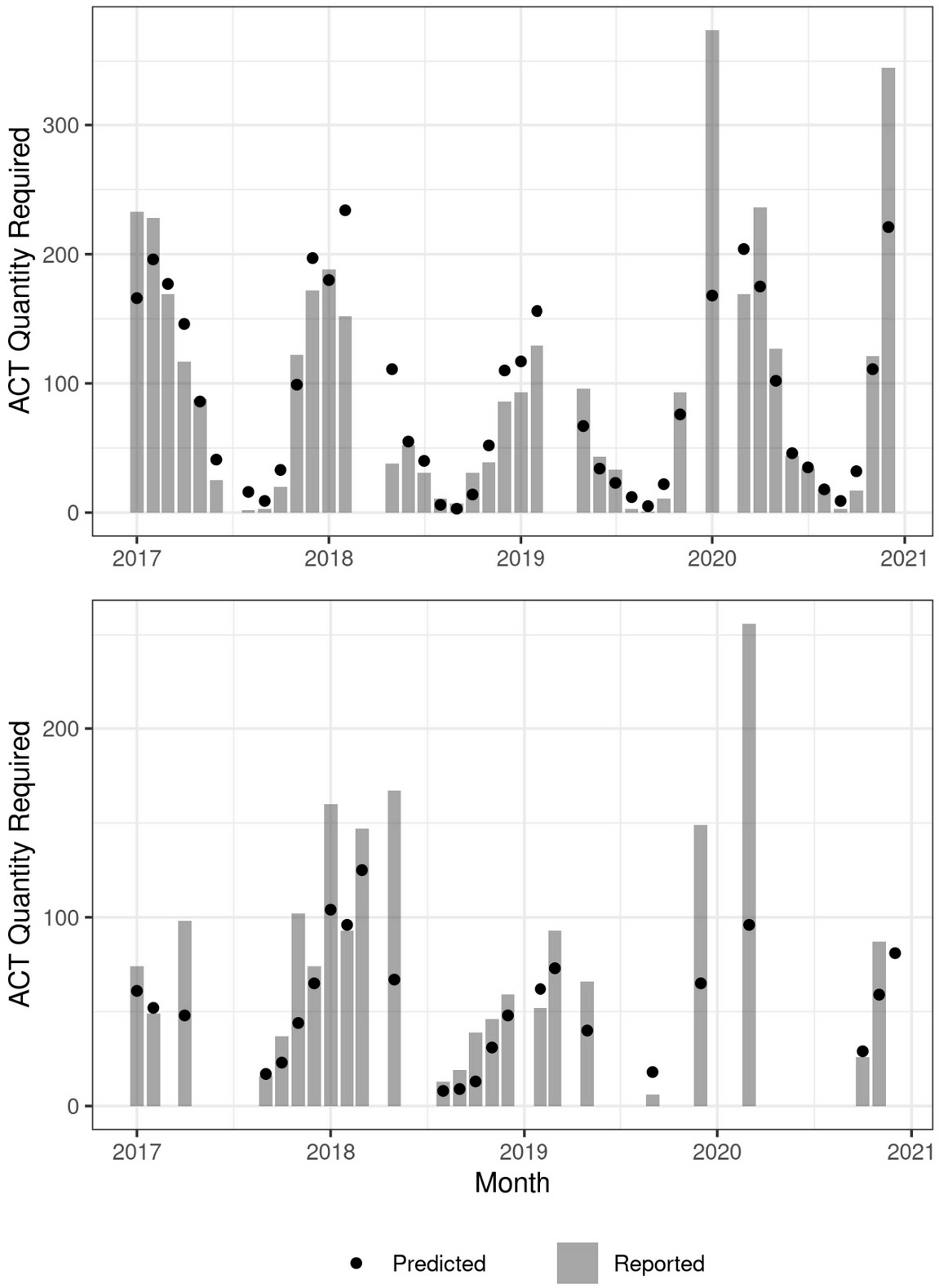
The ability of the SMALLER MEWS to accurately reproduce historical ACT requirements differed by PHC. This figure demonstrates two example PHCs where the SMALLER MEWS predicted the required ACT within 0.7% (top) and 51.2% (bottom) of the reported annual requirement. Months with missing reported ACT requirements are not plotted.

While the objective of this model was to predict future malaria cases based on established relationships with environmental variables, and not draw inference about these relationships, this does not preclude the exploration of these relationships established via the statistical model. We found that the time since the previous LLIN distribution was strongly associated with higher incidences of malaria (Fig. 4, Table S5.2). Fokontany with higher socio-economic levels tended to have a lower incidence of malaria (Fig. 4, Table S5.2). The majority of the 3-month lagged dynamic environmental variables were significantly associated with malaria incidence (Fig 4, Table S5.2). Malaria incidence was strongly associated with temperature and MNDWI at the village-level (Table S5.2, Fig. 4). It was also associated with EVI at the village-level, rainfall, and Rice Indices 2 (vegetation dynamics) and 3 (seasonal anomalies), although these coefficients were smaller than for temperature and MNDWI (Table S5.2, Fig. 4).

**Figure 4.**
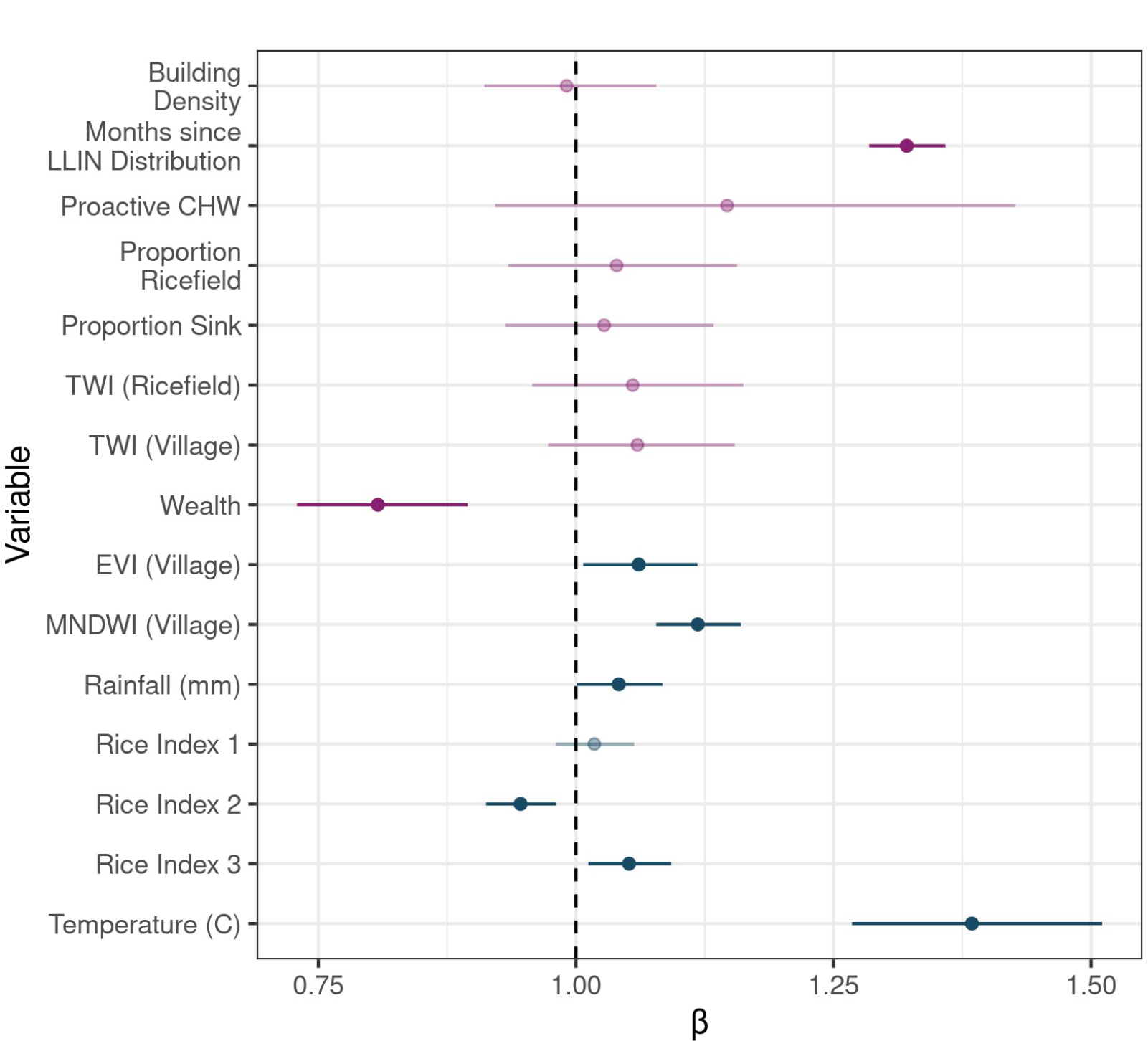
Model coefficients of the primary INLA model. Static variables are colored in purple and dynamic variables, which are lagged by 3 months and updated monthly, are colored in blue. Points represent the median value of the coefficient and error bars the 95% confidence intervals (CI). Those points whose CI overlaps 1, and therefore do not exhibit a statistically significant relationship with malaria incidence, are represented in faded colors.

### SMALLER MEWS Web Application

We integrated the INLA model within the automated workflow to predict malaria incidence three months in advance for each fokontany. These predictions are accessible via a dashboard-style web application which provides hyper-local information relevant to program managers and personnel working across the health system in Ifanadiana district. The landing page of the application provides an overview of malaria burden across the district over the next three months (Fig. 5). This includes alert buttons displaying four key indicators for district health personnel: the total number of cases, the total incidence, the malaria burden compared to the prior year, and the number of health clinics expected to receive more cases than the prior year. Also on this landing page is a map of the incidence in the district, displayed at the spatial scale of the community health catchment. Selecting a fokontany on this map opens a window which displays a time series of the forecasted malaria incidence, with historical time series included for context. These data can be explored further via a table that displays these values by month and fokontany. The table can be subset interactively and all of the data are available for download as a csv file.

One objective of this dashboard was to address health actors’ concerns regarding disruptions to diagnostic and medical stocks. We therefore added modules to help inform the risk of stock disruptions for each major PHC and the expected number of cases requiring treatment at the community health catchment. The module displaying stockout risk for each PHC compares the predicted number of cases expected to seek care at a PHC with the historical ACT use over the past two years. We also estimated the number of malaria cases not captured by PHCs and remaining at the community health level, presented in a seperate module. These cases could seek care at a community health site, be identified during proactive community health visits, or remain untreated. These functionalities are meant to provide granular information for district managers and health workers for context-specific decision-making.

[Figure removed because it contains text in French.]

Figure 5. **Screenshot of the landing page of the SMALLER MEWS web application.** The side panel serves to navigate between the pages of the site, including a page to download the data and explore predicted needs at the community and primary health center level. The web application can be accessed at https://smaller.pivot-dashboard.org/.

## DISCUSSION

Advances in disease analytics and forecasting, coupled with the increased availability of timely health data and fine resolution remotely-sensed satellite information, promise a new era of precision public health which will allow the delivery of “the right intervention to the right population at the right time” (Khoury et al. 2016). Yet, there is currently an important gap between the spatio-temporal scales at which these tools are available, and the much finer scales necessary to inform local program implementation for improving disease surveillance and control. We developed a hyper-local malaria early warning system (MEWS), SMALLER, for use by a health system strengthening program serving a rural health district in southeastern Madagascar. SMALLER combines fine-resolution information on static hydrological and socio-demographics with fine-resolution dynamic data derived from satellite imagery and climate models that are updated on a monthly frequency. SMALLER forecasted malaria incidence at the community scale (a village or group of villages) up to three months in advance, improving correlations with out-of-sample data by nearly 500% compared to a null model. When used to inform stock requirements, it predicted historical ACT requirements within 50 cases of the reported requirement over 68% of the time. Integrated into a web application, SMALLER provides real-time access for local health actors to the MEWS predictions to aid in context-specific decision making.

SMALLER contributes to a rapidly growing ecosystem of disease forecasting tools developed to aid in decision making. A recent review found that of the 37 existing tools used for the modeling of climate-sensitive infectious diseases, 16 of them focused exclusively on malaria (Ryan et al. 2023). The sensitivity of malaria to climate and environmental variables makes it an ideal candidate for disease forecasting efforts (Wimberly et al. 2021). Indeed, we found that ecological variables related to temperature and vegetation dynamics were strongly associated with malaria incidence, in agreement with past work (Arisco et al. 2020, Rakotoarison et al. 2020, Pourtois et al. 2023, Epstein et al. 2023). Interestingly, after accounting for seasonal dynamics via a cyclical temporal structure, MNDWI was more strongly associated with malaria incidence than precipitation. This variable was collected at a finer spatial resolution (10m vs. 10 km) that represented local standing water dynamics.

Weather stations are rare across the African continent, resulting in downscaled gridded precipitation data of low-quality (Dinku et al. 2022). This suggests that indices derived from satellite imagery may be more useful than those from coarser precipitation models for malaria prediction at local scales in areas with poor weather station coverage.

A feature unique to the SMALLER MEWS is its ability to integrate disease forecasts with information on historical stock quantities and disruptions. The ability of stock-outs to hinder progress towards malaria elimination has been highlighted since the introduction of ACTs nearly fifteen years ago (Kangwana et al. 2009, The PLoS Medicine Editors 2009). A stock-out not only prevents an individual patient’s treatment, but can increase healthcare costs when patients must seek treatment at private facilities (Mikkelsen-Lopez et al. 2013). Lower or delayed treatment rates can, in turn, allow for increased severity, onward transmission, and higher population-level prevalence rates (Challenger et al. 2019). A multinational study of eight sub-Saharan African countries found that, in contrast to the other seven countries in the region, Madagascar experienced a decrease in malaria diagnostic availability in public health facilities from 2010 to 2015 (Akulayi et al. 2017). Both RDTs and ACTs are delivered to public health facilities via a “pull system”, where health facility managers manually fill in quarterly orders for medicines and supplies, which are delivered from the capital to district depots and eventually individual public facilities’ pharmacies. Per national policy, the quantity requested is a function of the amount of materials dispensed during the prior quarter. Given the strong seasonality of malaria in Ifanadiana, particularly its exponential growth between the months of October - January, basing future needs on recent use can result in both under- and over-estimation of the quantities needed, depending on the season. While promising, the SMALLER MEWS was only able to predict stock needs within 50 cases of reported needs less than 70% of the time. Health-seeking behaviors and stock requirements are determined by complex economic and behavioral processes that govern the health system, and therefore fluctuate greatly from month to month. Particularly at the hyper-local scale, relatively small stochastic events, such as a family-level outbreak or a several-day disruption of a transportation route, can drastically influence the number of cases seen at the health center that month. SMALLER MEWS was unable to capture these months with uncharacteristically high or low requirements for the season, particularly in more recent years during the COVID-19 pandemic, but predictions could be improved through the integration of more recent health system data to inform the back-calculation.

Limitations to the scaling-up of SMALLER are primarily related to data constraints. An ideal MEWS would be directly connected to an electronic HMIS, such as DHIS2 in the case of Madagascar and many other countries, to facilitate the timely incorporation of the most recent disease and stock data. However, HMIS data are often reported at the scale of the PHC catchment, which comprises dozens of villages across hundreds of square kilometers, and rarely at spatial scales relevant to local targeting by community health workers and mobile teams. Handwritten PHC registries were manually digitized to obtain the granular dataset needed to train the statistical model in the SMALLER MEWS, a resource- and time-intensive process that is not scalable at a national level for routine surveillance.

However, the expansion of eHMIS systems and mobile technology such as commCare or DHIS2 tracker, which include information on patient residences, will make the integration of HMIS data into a national, highly granular MEWS more feasible in the future. Indeed, an electronic data collection program at PHCs was established in the district of Ifanadiana in October 2023, and will be integrated into a MEWS in the coming years as it expands to cover the entire district. Environmental data can also be limiting, not necessarily due to their availability but rather to the computational and technical resources required to access and process datasets at local scales (Wimberly et al. 2021). For example, a day of Sentinel-2 imagery for the country of Madagascar contains over 60 images, each 500-700 MB in size. Downloading this volume of data would be difficult with limited internet connectivity and the treatment and processing of the images would require a high-processing computer, given their quantity and size. Services which process satellite imagery on a remote server, such as Google Earth Engine, AWS, or MOSAIKS (Rolf et al. 2021), provide access to processed data without downloading the images themselves, but still require paid accounts and geospatial expertise to use. The on-going push for building disease forecasting capacity among public health actors and organizations will require further investment to make these data available in regions with low connectivity and computational resources for a successful and timely integration into health information systems.

## CONCLUSIONS

While recent advancements in data availability and statistical modeling have led to rapid growth in the development of MEWSs, few have been developed at the community scale required by certain public health interventions. We combined fine-resolution routine health data and environmental data from satellite imagery into a MEWS capable of generating predictions at a hyper-local scale. Cross-validation exercises revealed that the statistical model on which the MEWS was based had high predictive capacity across both space and time when applied to out-of-sample datasets. However, the SMALLER MEWS exhibited lower performance when predictions were used to estimate monthly medical stock requirements. This highlights a limitation of both the SMALLER MEWS and MEWSs in general regarding their ability to adequately account for the complex processes determining medical stock requirements and use. Future work should focus on how to integrate the already highly-predictive environmentally-driven MEWS into more complex models of health system functioning and processes to better understand and predict resource needs.

## DECLARATIONS

### Ethics Approval and Consent to Participate

Use of aggregate monthly consultation counts from PHC registers for this study was authorized by the Madagascar National Ethics Committee and the Medical Inspector of Ifanadiana district. It was deemed non-human subjects research by Harvard University’s Institutional Review Board. The IHOPE longitudinal survey implemented informed consent procedures approved by the Madagascar National Ethics Committee and the Madagascar Institute of Statistics. This included obtaining informed consent from all subjects or their legal guardians. Household-level de-identified data from the IHOPE survey were provided to the authors for the current study.

### Consent for Publication

Not Applicable

### Availability of Data and Materials

Data and code needed to reproduce these analyses can be found in a figshare repository (private link:https://figshare.com/s/35bb8aecbf5191f8dbba, public DOI to be shared upon publication). The web application’s source code can be found at https://gitlab.com/pivot-sci-apps/smaller-backend and https://gitlab.com/pivot-sci-apps/smaller-shiny.

### Authors’ Contributions

MVE, MHB, BR, and AG conceptualized the study. MVE, FAI, MR, VH, CR, TC, ED, BR, EM, FAR, BR and AG contributed to designing the methodologies concerning data collection and analysis. MVE, FAI, MR, VH, CR, TC, ED, BR and AG collected the data and MVE, FAI, MR, VH, CR, TC, ED, BR, EM, FAR, BR, OR and AG conducted analyses and interpreted results. Project supervision was provided by FAI, VH, CR, MHB, BR, BR, OR and AG. The manuscript was drafted by MVE and AG and all authors contributed critically to the drafts and gave final approval for publication.

## Data Availability

Data and code are available upon request to the authors and will be made publicly available online upon publication.

## Acknowledgements

We would like to thank the health professionals of Ifanadiana, who serve their patients while aiding in the collection of vital health data. We would like to acknowledge Ann Miller and the research team at INSTAT, who implement the IHOPE survey. This work would not be possible without the Pivot data collection team. We would also like to thank the Pivot clinical and programmatic teams for their feedback on this project and the Pivot IT team for their logistical support. We would also like to thank Thomas Germain, Pascal Mouquet, and the research group at SEAS-OI for deploying an automated Sen2Chain extraction workflow for our use.

