## Supplemental Materials for "Increasing the resolution of malaria early warning systems for use by local health actors"

Table of Contents

1. Creating Rice Indices
2. Configuring the ZERO-G adjustment for the study zone
3. Comparing Non-linear vs. Linear Model Performance
4. Supplemental Figures
5. Supplemental Tables

### S1. Creating Rice Indices via Principal Components Analysis

We used principal component analysis to create indices of rice field dynamics from EVI, MNDWI, and GAO-NDWI extracted from rice fields zones. First, we estimated the difference in each indicator at the rice field level from the indicator extracted at a buffer of 1 km surrounding village zones (Δvill-rice) to categorize dynamics specific to rice field environments. Second, we transformed the rice-field level indicators (EVI, MNDWI, and GAO-NDWI) into seasonal anomalies by standardizing each indicator within each calendar month (e.g. January, February, etc.) following Kaul et al. (2018). This seasonal anomaly represents how conditions differed in that month compared to the same calendar month of different years. Third, we included the original three environmental indicators extracted and average at the major rice field zones. We used all three forms of the three indicators ( Δvill-rice, seasonal anomalies, and original mean; 9 variables total) in the PCA, after having centered and scaled each one. We then selected the first three components, which contained over 73% of the overall variance, to represent three indices of rice field dynamics (Table S1.1, Table S1.2). Rice Index 1 was strongly influenced by all three forms of the MNDWI indicator, and represented the amount of standing water in rice fields. Rice Index 2 was more strongly associated with EVI and NDWI-GAO and represented the vegetation dynamics of the rice fields. Rice Index 3 was most strongly associated with anomalies in the vegetation indices and represented anomalies in vegetation phenology and the timing of the agricultural season.

|  | **PC1** | **PC2** | **PC3** | **PC4** |
| --- | --- | --- | --- | --- |
| **Standard Deviation** | 1.708 | 1.560 | 1.086 | 0.921 |
| **Proportion of Variance** | 0.324 | 0.284 | 0.131 | 0.094 |
| **Cumulative Proportion** | 0.324 | 0.609 | 0.740 | 0.834 |

**Table S1.1.** Proportion of variance explained by the first four components of the PCA used to create the rice indices.

|  | **Dim. 1** | **Dim. 2** | **Dim. 3** |
| --- | --- | --- | --- |
| **MNDWI** | 30.81 | 0.21 | 1.05 |
| **EVI** | 0.98 | 31.59 | 3.56 |
| **NDWI-GAO** | 6.94 | 16.82 | 3.11 |
| **ΔMNDWI** | 28.95 | 1.03 | 0.96 |
| **ΔEVI** | 8.17 | 17.55 | 4.59 |
| **ΔGAO** | 9.9 | 10.03 | 14.54 |
| **MNDWI Anomalies** | 10.24 | 0 | 9.63 |
| **EVI Anomalies** | 0.2 | 12.49 | 25.42 |
| **GAO Anomalies** | 3.82 | 10.28 | 37.15 |

**Table S1.2.** Contribution of each variable to the first three components.

|  | **Dim. 1** | **Dim. 2** | **Dim. 3** |
| --- | --- | --- | --- |
| **MNDWI** | 0.948 | -0.073 | -0.111 |
| **EVI** | -0.169 | 0.899 | -0.205 |
| **NDWI-GAO** | 0.45 | 0.656 | -0.192 |
| **ΔMNDWI** | -0.919 | 0.163 | 0.106 |
| **ΔEVI** | 0.488 | -0.67 | 0.233 |
| **ΔGAO** | -0.537 | -0.507 | 0.414 |
| **MNDWI Anomalies** | 0.547 | 0.003 | 0.337 |
| **EVI Anomalies** | -0.076 | 0.565 | 0.548 |
| **GAO Anomalies** | 0.334 | 0.513 | 0.662 |

**Table S1.3** Coordinates of each variable for the first three PCA components.

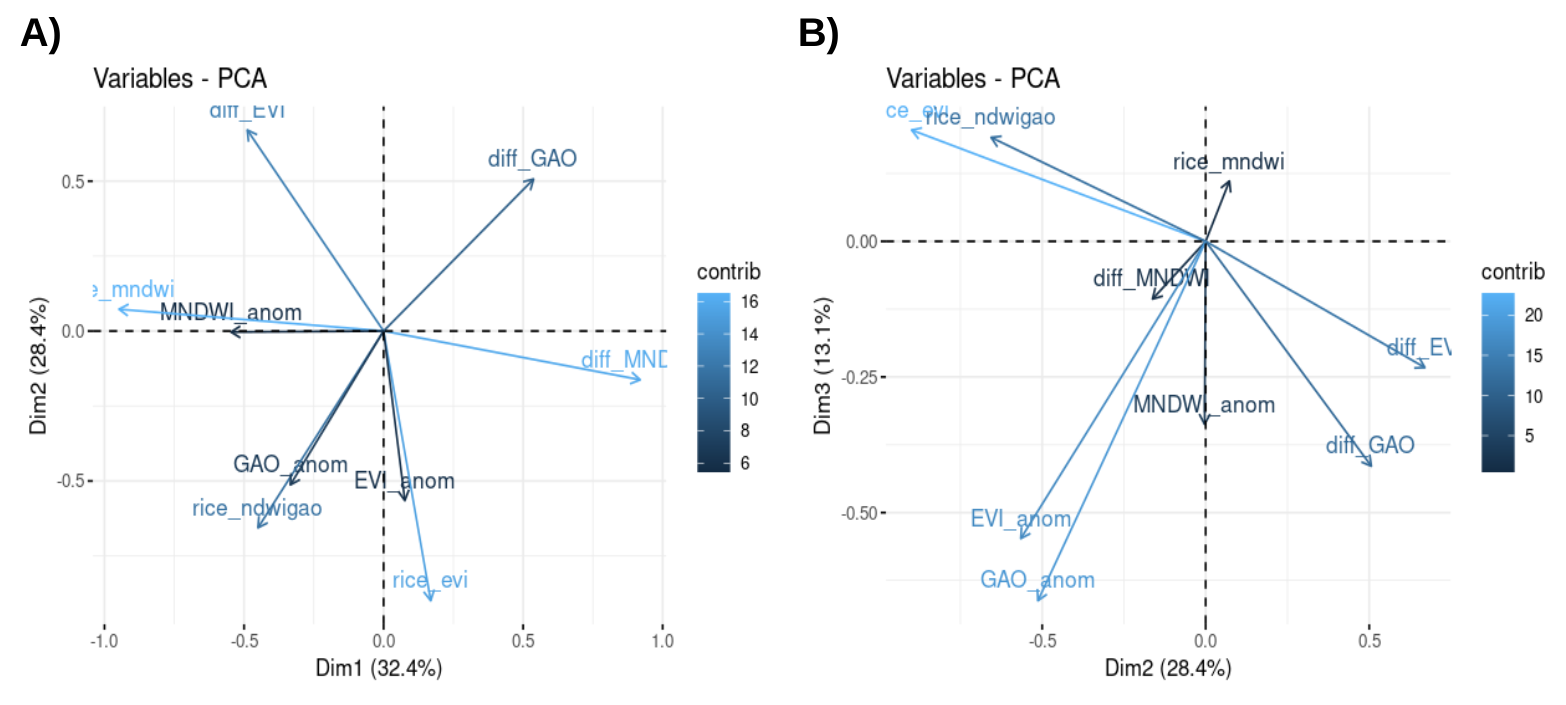

**Figure S1.1.** Plot of PCA Loadings for components 1 and 2 (A) and components 2 and 3 (B).

### S2. Configuring the ZERO-G adjustment for the study zone

We applied a zero-adjusted gravity-model estimator (ZERO-G, Evans et al. 2023) to adjust the malaria case data for biases due to geographic barriers, financial barriers, and differing levels of health system support across space and time. We applied the following configurations to the ZERO-G algorithm so that it better conformed to our study system and period:

- Limited the linear trend to be positive, to account for increasing access to the health system over time due to the ongoing health-system strengthening intervention
- Included a “service” of PHCs that represented the COVID-19 pandemic as a binary variable, to account for reduced use of PHCs between March 2020 - July 2021. This allowed the “mass” of a clinic to be reduced during this time period.
- Did not include fokontany missing more than 50% of data or with more than 50% months reporting zero consultations in our estimation of parameters for estimating healthcare access
- Identified zero’s to be imputed via local context. Only those zeros outside of the HSS intervention during the malaria high season (Nov-April) were imputed

### S3. Comparing Non-Linear vs. Linear Model Fit

Many studies have found the relationship between malaria burdens and environmental variables to be non-linear (Okiring et al. 2021, Odhiambo et al. 2020, Zinzser et al. 2012). This is particularly true for climatic variables such as temperature and rainfall, which exhibit unimodal relationships with malaria where malaria burdens decrease at the extremes of the range (Mordecai et al. 2019, Paiijmans et al. 2007). We therefore explored implementing a non-linear model via penalized smoothing splines. Penalized smoothing splines add a penalty to the model to prevent the splines from overfitting the data. We included a penalized smoothing spline for each variable and compared this model to our original linear model described in the main text. We found little difference in model performance or predictions between the two models (Fig. S2.1). Both models had a Spearman’s correlation of 0.64 with the true data and an RMSE of 61.5. However, the runtime of the non-linear model was nearly 100x longer than the linear model. We therefore chose the more parsimonious linear model for the application.

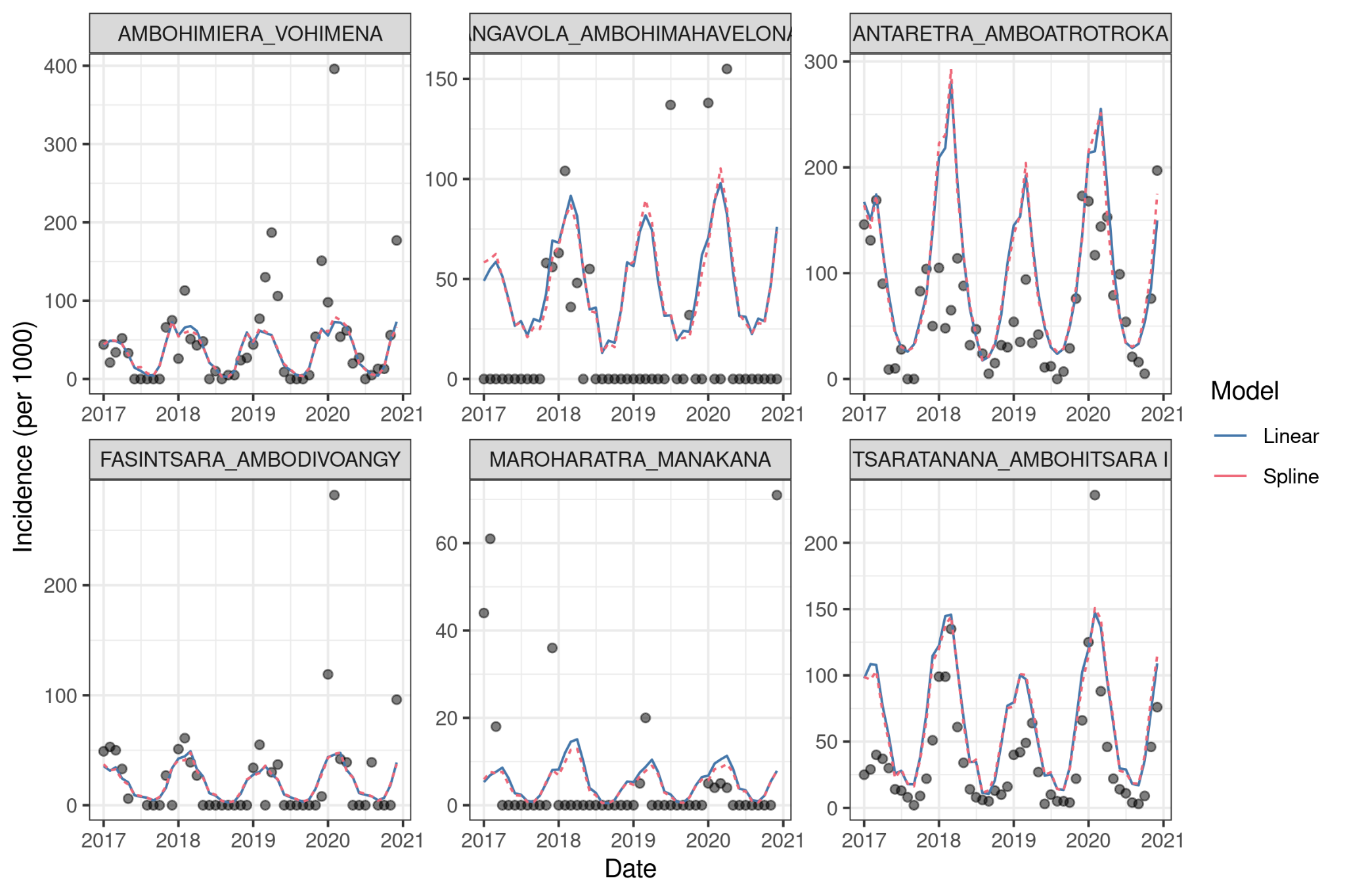

**Figure S2.1. Predictions did not differ between the linear and non-linear penalized spline model.** Predicted incidence rates are plotted for six randomly sampled fokontany, with the true rates shown in black points.

### S4. Supplemental Figures

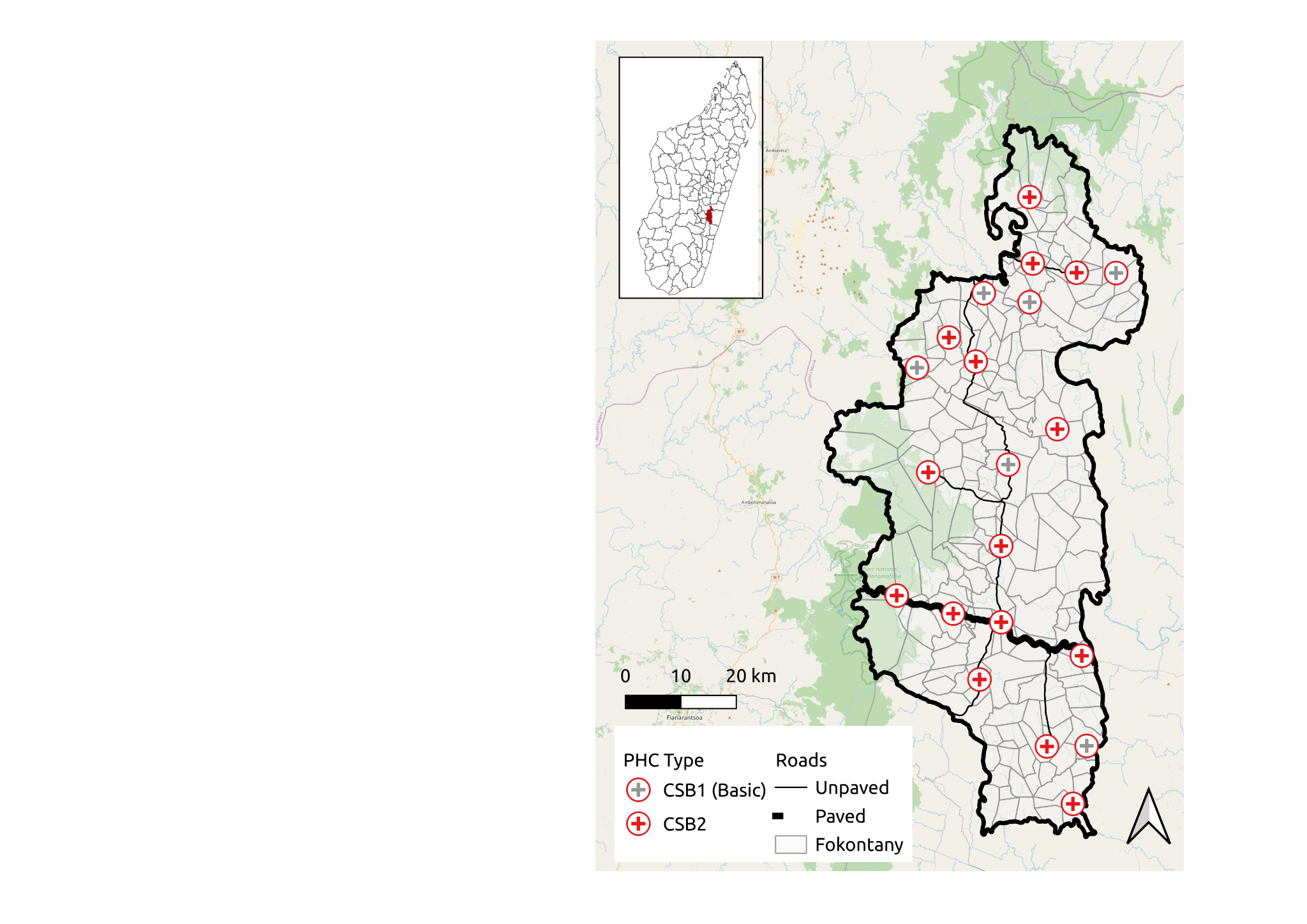

**Figure S4.1. Map of study area (District of Ifanadiana, Vatovavy, Madagascar).** Inset panel locates the district with the country. Background map is sourced from OpenStreetMap.

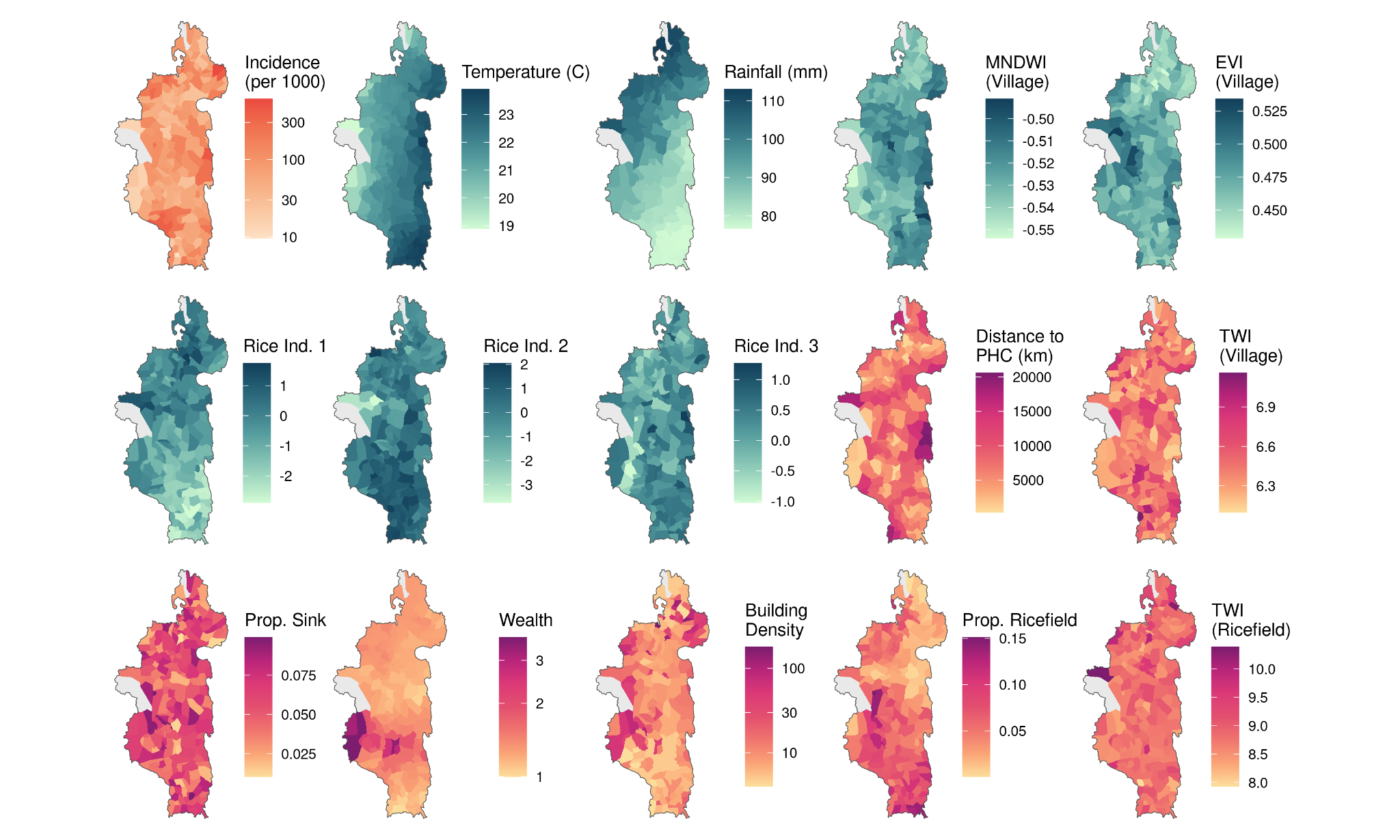

**Figure S4.2. Maps of malaria incidence and covariates used in the forecasting model at the fokontany level.** Dynamic variables are colored in teal and the average value during the year 2020 is shown. Static variables are plotted in red.

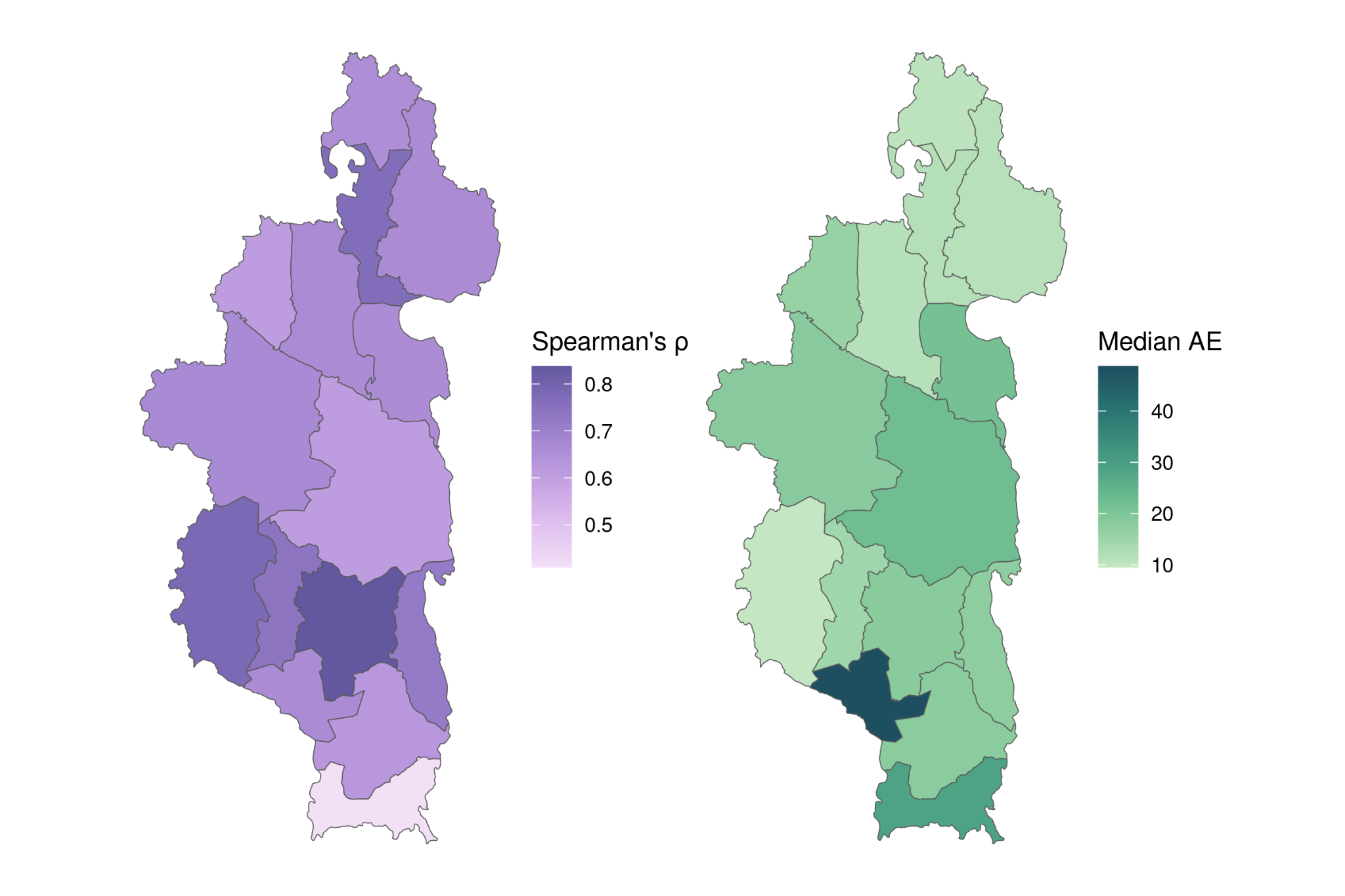

**S4.3. Performance metrics by each spatial block (commune) used as out-of-sample data in the cross-validation procedure.** Median AE represents the median absolute error of between the predicted and observed incidence rates when each commune was out-of-sample, with lower error rates representing higher accuracy.

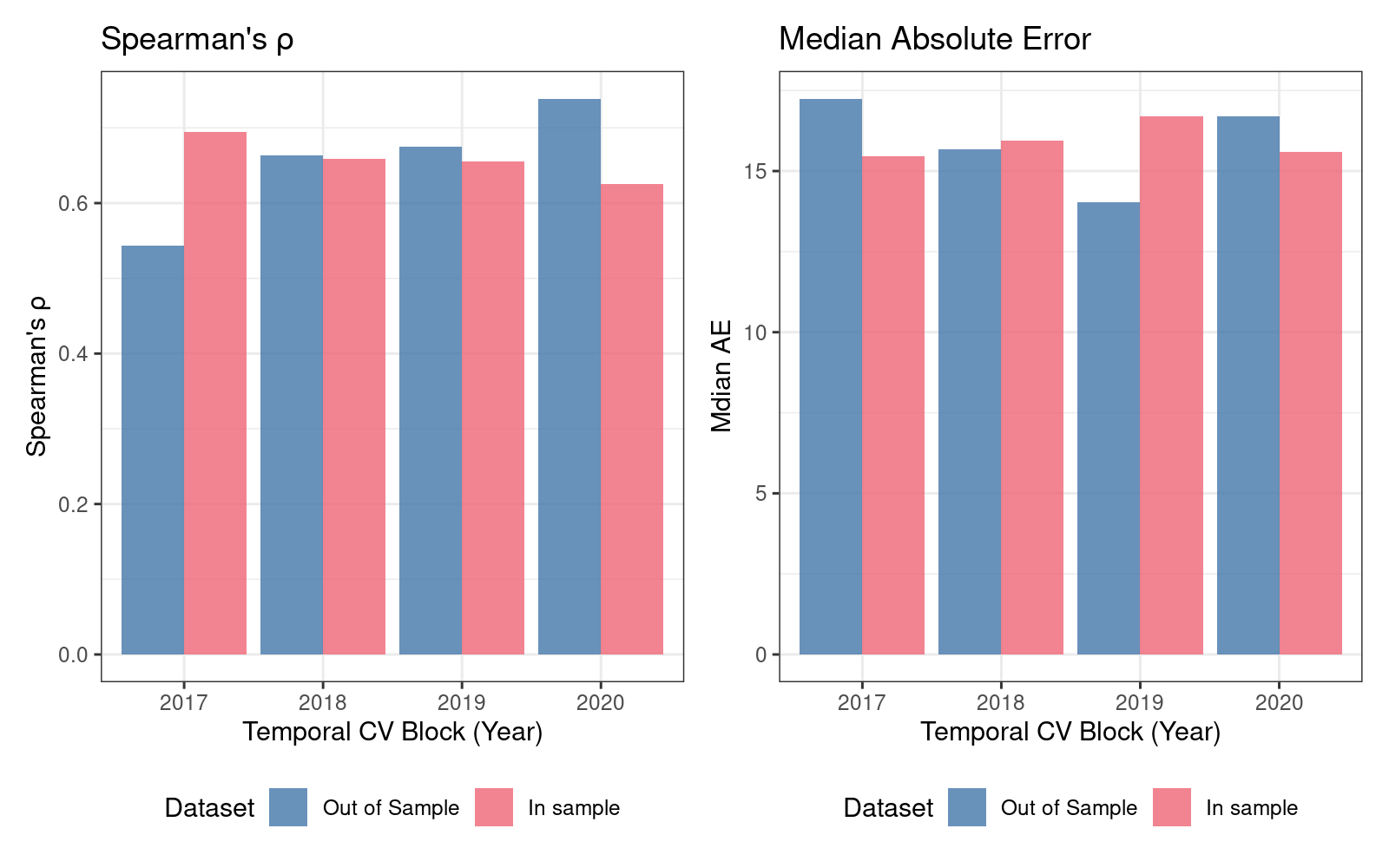

**S4.4. Performance of the model applied to temporal cross-validation by year, with each year representing an out-of-sample cross-validation block.**  Median AE represents the median absolute error of between the predicted and observed incidence rates, with lower error rates representing higher accuracy.

### S5. Supplemental Tables

**Table S5.1 Details of predictor variables used in the full INLA model.**

| **Variable** | **Temporal Frequency** | **Spatial Resolution** | **Lag** | **Source** |
| --- | --- | --- | --- | --- |
| Building Density | Static | Fokontany | NA | OpenStreetMap |
| Months since LLIN Distribution | Monthly | Fokontany | 0 months | MMoPH |
| Proactive CHW | Monthly | Fokontany | 0 months | Pivot |
| Proportion Ricefield | Static | 10m (Village Extraction) | NA | Pivot |
| Proportion Sink | Static | 30m (Village Extraction) | NA | SRTM |
| TWI (Ricefield) | Static | 30m (Village Extraction) | NA | SRTM |
| TWI (Village) | Static | 30m (Ricefield Extraction) | NA | SRTM |
| Wealth | Static | Fokontany | NA | Pivot |
| EVI (Village) | Monthly | 10m (Village Extraction) | 3 months | Sentinel-2 |
| MNDWI (Village) | Monthly | 10m (Village Extraction) | 3 months | Sentinel-2 |
| Precipitation | Monthly | 0.1 degree (Village Extraction) | 3 months | ARC2 |
| Rice Index 1 | Monthly | 10m (Ricefield Extraction) | 3 months | Sentinel-2 |
| Rice Index 2 | Monthly | 10m (Ricefield Extraction) | 3 months | Sentinel-2 |
| Rice Index 3 | Monthly | 10m (Ricefield Extraction) | 3 months | Sentinel-2 |
| Land Surface Temperature | Monthly | 1 km (Village Extraction) | 3 months | MODIS |

**Table S5.2. Coefficients of fixed effects in full INLA model.**

| **Variable** | **Lower 95% CI** | **Median** | **Upper 95% CI** |
| --- | --- | --- | --- |
| Months since LLIN Distribution | 1.285 | 1.321 | 1.359 |
| Proactive CHW | 0.922 | 1.147 | 1.427 |
| Proportion Ricefield | 0.934 | 1.039 | 1.156 |
| Proportion Sink | 0.931 | 1.027 | 1.134 |
| TWI (Ricefield) | 0.958 | 1.055 | 1.162 |
| TWI (Village) | 0.973 | 1.06 | 1.154 |
| Wealth | 0.729 | 0.808 | 0.895 |
| Building Density | 0.911 | 0.991 | 1.078 |
| Temperature (C) | 1.268 | 1.384 | 1.511 |
| Rainfall (mm) | 1.001 | 1.042 | 1.084 |
| MNDWI (Village) | 1.078 | 1.118 | 1.16 |
| EVI (Village) | 1.007 | 1.061 | 1.118 |
| Rice Index 1 | 0.981 | 1.018 | 1.057 |
| Rice Index 2 | 0.913 | 0.946 | 0.981 |
| Rice Index 3 | 1.012 | 1.052 | 1.093 |
